# Effects of Robotic-Assisted Gait at Different Levels of Guidance and Body Weight Support on Lower Limb Joint Angles, Angular Velocity, and Inter-Joint Coordination

**DOI:** 10.1101/2022.05.16.22275104

**Authors:** Yosra Cherni, Yoann Blache, Mickaël Begon, Laurent Ballaz, Fabien Dal Maso

**Affiliations:** Centre interdisciplinaire de Recherche en Réadaptation et Intégration Sociale, Québec, QC, Canada; Département de réadaptation, faculté de Médecine, Université Laval, Québec city, QC, Canada; Centre de Recherche et d’Innovation sur le Sport, Université de Lyon, Lyon, France; Laboratoire de Simulation et Modélisation du Mouvement, École de kinésiologie et des sciences de l’activité physique, Université de Montréal, Montréal, QC, Canada; Centre de recherche du CHU Ste Justine, Montréal, QC, Canada; Département des sciences de l’activité physique, Université du Québec à Montréal, Montréal, QC, Canada; Centre interdisciplinaire sur le cerveau et l’apprentissage, Université de Montréal, QC, Canada

**Keywords:** Lokomat, Treadmill, Gait pattern, Angular velocity, Continuous Relative Phase, Statistical parametric mapping

## Abstract

The Lokomat provides task-oriented therapy for patients with gait disorders. This robotic technology drives the lower limbs in the sagittal plane. However, unconstrained gait involves motions also in the coronal and transverse planes. This study aimed to compare the Lokomat with Treadmill gait through 3D-joint kinematics and inter-joint coordination. Lower limb kinematics was recorded in 18 healthy participants who walked at 3 km/h on a Treadmill or in a Lokomat with nine combinations of Guidance (30, 50, 70%) and body-weight-support (30, 50,70%). Compared to Treadmill, the Lokomat altered pelvis rotation, decreased pelvis obliquity and hip adduction, and increased ankle rotation. Moreover, the Lokomat resulted in a significantly slower velocity at the hip, knee, and ankle flexion compared to the treadmill condition. Moderate to strong correlations were observed between the Treadmill and Lokomat conditions in terms of inter-joint coordination between hip-knee (r=0.67-0.91), hip-ankle (r=0.66-0.85), and knee-ankle (r=0.90-0.95). In conclusion, this study showed that some gait determinants such as pelvis obliquity and rotation, and hip adduction are altered when walking with Lokomat in comparison to Treadmill. Kinematic deviations induced by the Lokomat were most prominent at high levels of body-weight-support. Interestingly, different levels of Guidance did not affect gait kinematics.

## 1 Introduction

Over the past two decades, robotic-assisted gait orthoses have emerged to provide task-oriented and mass-practice therapy in individuals with neurological movement disorders (Nam et al., 2017; Swinnen et al., 2015b). These devices aim to drive patients’ lower limbs in a physiological gait pattern, limiting the strenuous and exhausting aspect of manual-assisted gait therapy. Patients can therefore benefit from longer interventions with more repetitions. The Lokomat is amongst the most used robotic-assisted gait orthoses in rehabilitation centers (Nam et al., 2017). Combined with conventional therapies, the Lokomat-based training provides better functional improvements than conventional therapies alone (Chung, 2017; Dundar et al., 2014). Understanding the gait biomechanics when individuals walk using the Lokomat is crucial to anticipate better the effect of Lokomat-based therapies (Aurich-Schuler et al., 2017) and to design optimal conventional therapy performed to complement Lokomat-based training.

The Lokomat assists patients to walk on a Treadmill using a harness that dynamically supports their body weight and bilateral motorized leg orthoses that drive their hips and knees in a predefined sagittal gait pattern. Both body weight support (BWS) and leg guidance are adjustable between 0% and 100%. In practice, 100% of BWS supports all the patient’s body weight; and 100% Guidance drives legs throughout the gait range of motion without effort. Conversely, low percentages of BWS and Guidance require a higher contribution of the patients to maintain their upright posture and gait pattern. The therapist needs to adjust Guidance and BWS settings, as well as leg orthoses range of motion to increase patients’ involvement (Fischer et al., 2015; Goldberg and Stanhope, 2013). There is some, albeit limited, evidence from clinical trials that Guidance and BWS Lokomat settings might be essential to increase the impact of this technology on the gait biomechanics in individuals with neuromotor disorders (Cherni et al., 2017; Park et al., 2019; Rodrigues et al., 2017). However, the existing literature has not yet sufficiently investigated how Lokomat settings influence gait.

Interestingly, although the Lokomat only drives patients’ lower limbs in the sagittal plane, individuals walking in the Lokomat showed translations and rotations in the coronal and transverse planes. However, the range of motion of these movements differed compared to Treadmill gait. Indeed, Lokomat gait significantly reduced thorax and pelvis lateral displacements compared to Treadmill gait (Swinnen et al., 2015a, 2014). Reduced thorax flexion and axial rotation were also evidenced during Lokomat gait compared to Treadmill gait (Swinnen et al., 2015a, 2014). Moreover, Lokomat settings also affect gait kinematics. An increase in BWS significantly reduced the amplitude of thorax flexion and rotation (Swinnen et al., 2015b). In contrast, an increase in Guidance did not affect gait (Swinnen et al., 2015b). Concerning lower-limb kinematics, Hidler et al., (2008) showed a greater hip and ankle range of motion in the sagittal plane during Lokomat gait compared to Treadmill gait. However, to the best of our knowledge, no study investigated leg kinematics in the coronal and transverse planes. Moreover, normative gait pattern also involves a precise rhythmic pattern so that joint angular velocities (Mentiplay et al., 2018) and inter-joint coordination, assessed by the continuous relative phase (CRP) (Chiu et al., 2015), are fundamental gait parameters. However, the latters have not yet been studied during the Lokomat gait.

The objective of this study was to compare joint angles, angular velocities, and coordination during non-assisted Treadmill gait and assisted Lokomat gait with various levels of BWS and Guidance. This study involved healthy individuals as many patients with gait disorders cannot walk in a normative gait without assistance (Aurich-Schuler et al., 2019; Swinnen et al., 2015b). As the Lokomat has been designed to drive the gait in the sagittal plane, it was hypothesized that joint kinematics and coordination in this plane would present strong similarities between the Treadmill and Lokomat conditions. On the opposite, it is expected that joint kinematics would significantly differ between Treadmill and Lokomat conditions in the coronal and transverse planes. Finally, as only BWS reduced thorax and pelvis kinematics (Swinnen et al., 2015a, 2015b), we expected high values of similarity scores between Treadmill and Lokomat conditions with low percentages of BWS.

## 2 Materials and methods

### 2.1 Participants

Eighteen healthy adults (9 females, age: 26.5 ± 4.7 years; height: 1.72 ± 0.09 m; mass: 66.1 ± 11.4 kg), all right-footed, volunteered to this study. Participants were excluded if they reported any lower limb and back injuries and/or orthopedic surgery at the time of the experiment or during the last year. All participants provided a written informed consent approved by the Ethics Committee of Sainte-Justine Hospital (#4049).

### 2.2 Instrumentation

#### Lokomat

The Lokomat Pro (**Figure 1A**) was used for this study.

**Figure 1.**
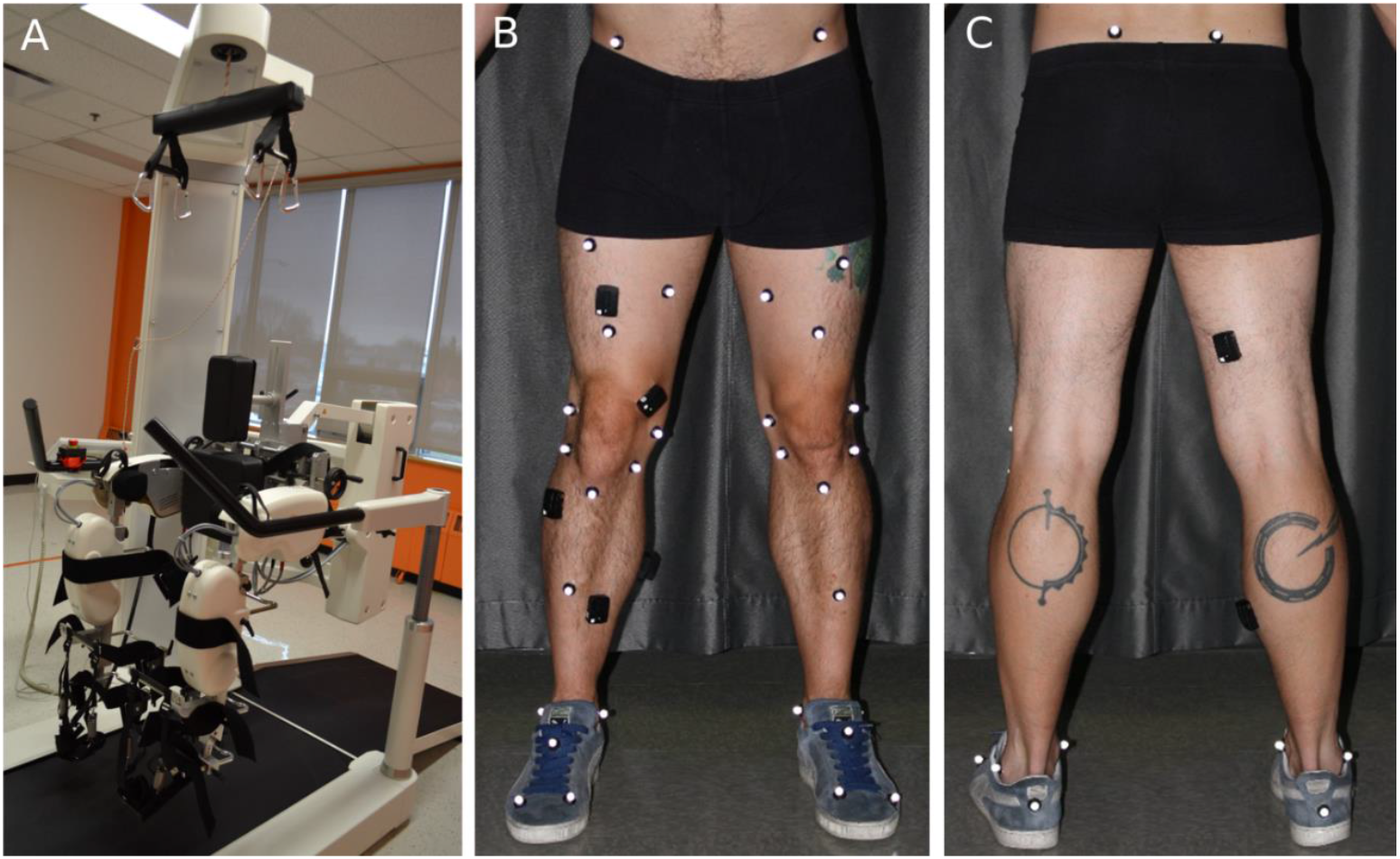
A) Lokomat. B) Front and C) back views of a participant equipped with reflective markers. Note: electromyography electrodes were put on the participants, but data were not used in the present study.

#### Kinematics

In line with International Society of Biomechanics recommendations(Wu et al., 2002) and considering the position of the Lokomat straps on the participants’ lower limbs, skin markers were positioned on the left and right foot (n=4), shank (n=6), thigh (n=5), and anterior superior iliac spine (n=1) (**Figure 1B-C**). In addition, during the static pose and functional movements, two markers were added on the posterior superior iliac spines (**Figure 1C**). These markers were not present during the experimental conditions as the Lokomat harness hid these bony landmarks. Marker trajectories were recorded using a 12-camera motion capture system (T-40 and T-20 Vicon cameras, Oxford Metrics Ltd., Oxford, UK) at a rate of 300 Hz.

### 2.3 Experimental procedures

#### 2.3.1 Familiarization session

Three to six days before the main experimental session, the exoskeleton was suited to the participants’ anthropometry (i.e., legs, cuffs and harness sizes, joint alignments). Then, participants were familiarized with the Lokomat gait during a 45-minute active walk. During this familiarization session, participants walked in the Lokomat at various speeds (1.5 to 3.0 km/h), BWS (20% to 80%), and Guidances (20% to 80%). An experimenter, the same across all participants, adjusted the hip and knee range of motion and the synchronization between the speed of the Treadmill and the feet to mimic normative gait.

#### 2.3.2 Main experimental session

##### Lokomat and Treadmill gait

The experimental test consisted of walking at 3.0 km.h^-1^ in two modalities, namely, Lokomat gait and Treadmill gait. A speed of 3.0 km.h^-1^ was chosen because the Lokomat best operates close to this speed(Hidler et al., 2008) despite self-selected healthy adult over-ground walking speed ranges between 4.5 and 4.9 km.h^-1^ (Himann et al., 1988). The session started with a 15-min re-familiarization with the Lokomat gait. For the Lokomat conditions, participants walked with three levels of BWS (30%, 50%, and 70%) and three levels of Guidance (30%, 50%, and 70%). These nine experimental conditionswere performed in a random order. These levels of BWS and Guidance match the range of Lokomat settings reported during therapy (Chang et al., 2012; Cherni et al., 2020; Mayr et al., 2007; Mazzoleni et al., 2011; Varoqui et al., 2014). Participants were asked to walk actively with the Lokomat. Each experimental condition lasted 30 seconds and was followed by a 1-minute passive gait. Then, for the Treadmill condition, participants walked on the Treadmill of the Lokomat without the harness and the robotic legs.

##### Static pose and functional movements

To personalize a multibody kinematic model, participants held a standing posture for a few seconds and performed hip and ankle flexion-extension, abduction-adduction, and circumduction, as well as knee flexion-extension movements. These recordings were required to functionally locate hip and ankle joint centers of rotation, and knee axes of rotation (Begon et al., 2007).

### 2.4 Data processing

#### Joint kinematics reconstruction

Hip and ankle centers of rotation and knee axes of rotation were determined using functional methods (Ehrig et al., 2007, 2006). Then, joint kinematics were obtained using an inverse kinematics procedure based on a nonlinear least-squares algorithm (Thouzé et al., 2016). The multibody kinematic model was composed of seven segments, namely, pelvis and left and right thigh, shank, and foot articulated by 20 degrees-of-freedom, namely, three rotations and three translations at the pelvis, three rotations at each hip, flexion-extension at each knee and three rotations at each ankle. Although the kinematic model computed joint angles bilaterally, only the kinematic of the right leg was reported for further analysis. Moreover, we did not consider hip rotation, and ankle abduction-adduction as these degrees-of-freedom presented large variability.

#### Angular velocities

The joint angular velocities were then calculated as the relative angular velocity between segments for the pelvis, hip, knee and ankle. Moreover, as the walking speed was constant for all participants and all conditions of the Lokomat and Treadmill, the stride rate was calculated to better interpret the results of angular velocities.

#### Continuous relative phase

The inter-joint coordination was computed between hip, knee, and ankle flexion-extension degrees-of-freedom using the continuous relative phase (CRP). The phase angle of each degree-of-freedom was computed using the Hilbert transform (Lamb and Stöckl, 2014) before calculating the hip-knee, hip-ankle, and knee-ankle CRP.

#### Heel strikes

Heel strikes frames were automatically determined according to the trajectory of the marker located on the heel using a custom-made script. Time histories of joint angles, joint angular velocities, and CRP were time-normalized between two heel strike frames.

### 2.5 Statistics

To assess the effect of each Lokomat condition on gait pattern, two statistical procedures were employed. Firstly, paired t-tests from the statistical parametric mapping package (Pataky et al., 2015) were used to compare time-histories of joint angles and velocities, and CRP between each Lokomat condition (n=9) and the Treadmill condition. The significance threshold was set to 0.05. As 153 comparisons were performed – (7 joint angles + 7 joint angular velocities + 3 CRP) × 9 Lokomat conditions–, *p*-values were corrected using a false discovery rate procedure (Benjamini and Hochberg, 1995) with a rate set to 10% (Parent et al., 2019). Differences only lasting more than 5% of the gait cycle were considered for analysis. In addition, paired t-tests were used to compare the stride rates between each Lokomat conditions and the Treadmill condition. Secondly, Pearson correlation analyses were used to assess the similarity of joint angles, angular velocities, and CRP between each Lokomat condition (n=9) and the Treadmill condition.(Aurich-Schuler et al., 2019) Correlations in the range of 0.0-0.3, 0.3-0.7, and 0.7-1.0 and their negative counterparts were considered as weak, moderate, and strong correlation, respectively (Ratner, 2009).

## 3 Results

For joint angles (**Figure 2**), joint angular velocities (**Figure 4**), and CRP (**Figure 5**), significant differences between the Lokomat and the Treadmill conditions lasted qualitatively longer for Lokomat conditions with 70% of BWS than for Lokomat conditions with 30% and 50% of BWS (see **Table S1** for corrected *p*-values and interval of significant differences). The following sections compare the Lokomat gait to the Treadmill gait in terms of joint angles, joint angular velocities, and CRP.

**Figure 2.**
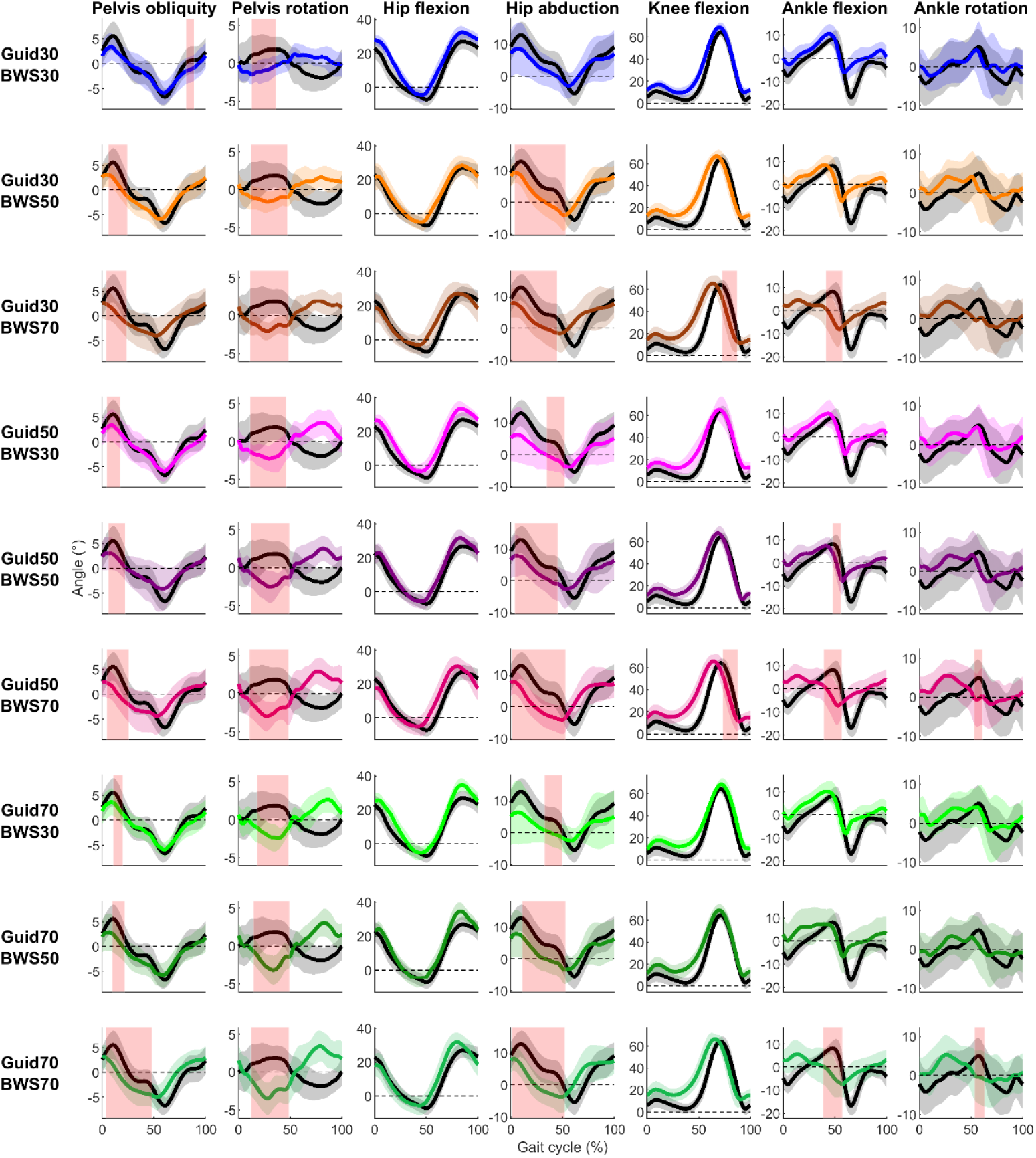
Participant’s mean (solid lines) ± standard deviation (shaded zones) joint angles during the Treadmill (black) and Lokomat (colored) conditions throughout the gait cycle for each degree of freedom (column) and Lokomat setting (row). Light red shaded zones report a significant difference between the Treadmill and Lokomat condition (FDR corrected *p*-values are reported in **Table S1**).

**Figure 3.**
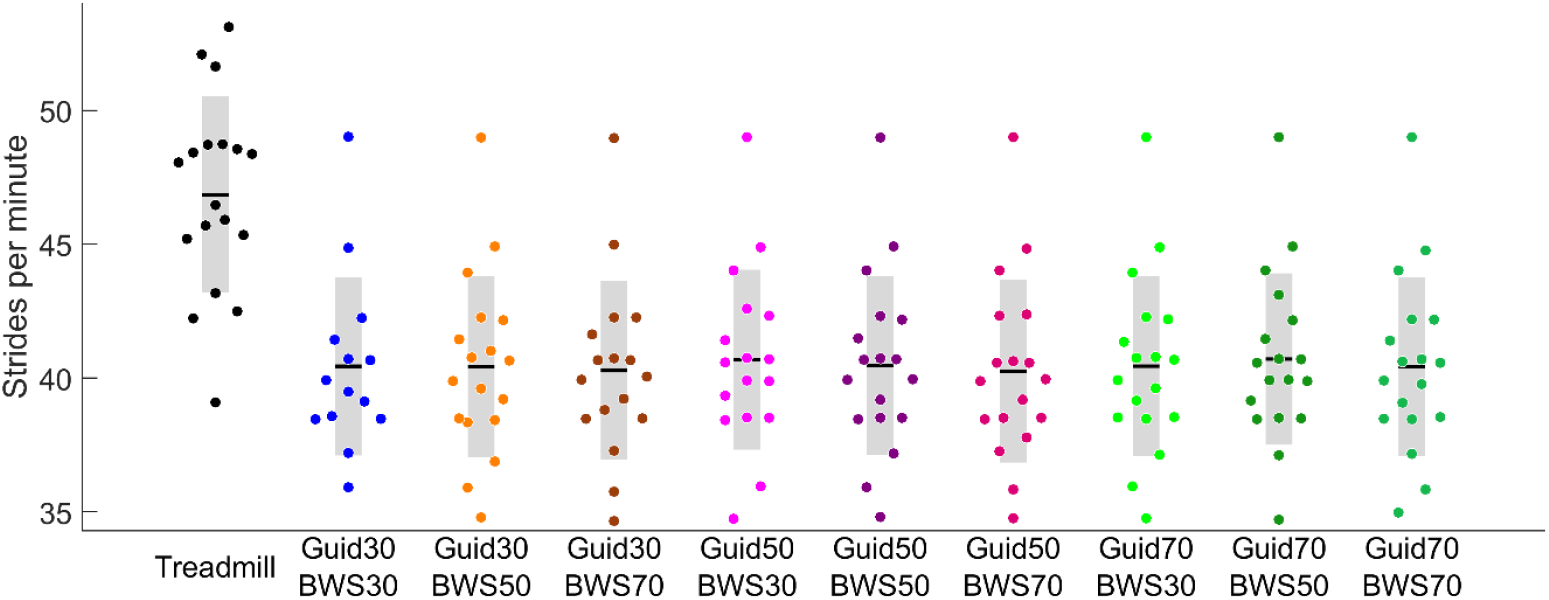
Stride rates in the Treadmill (black) and different Lokomat (colored) conditions.

**Figure 4.**
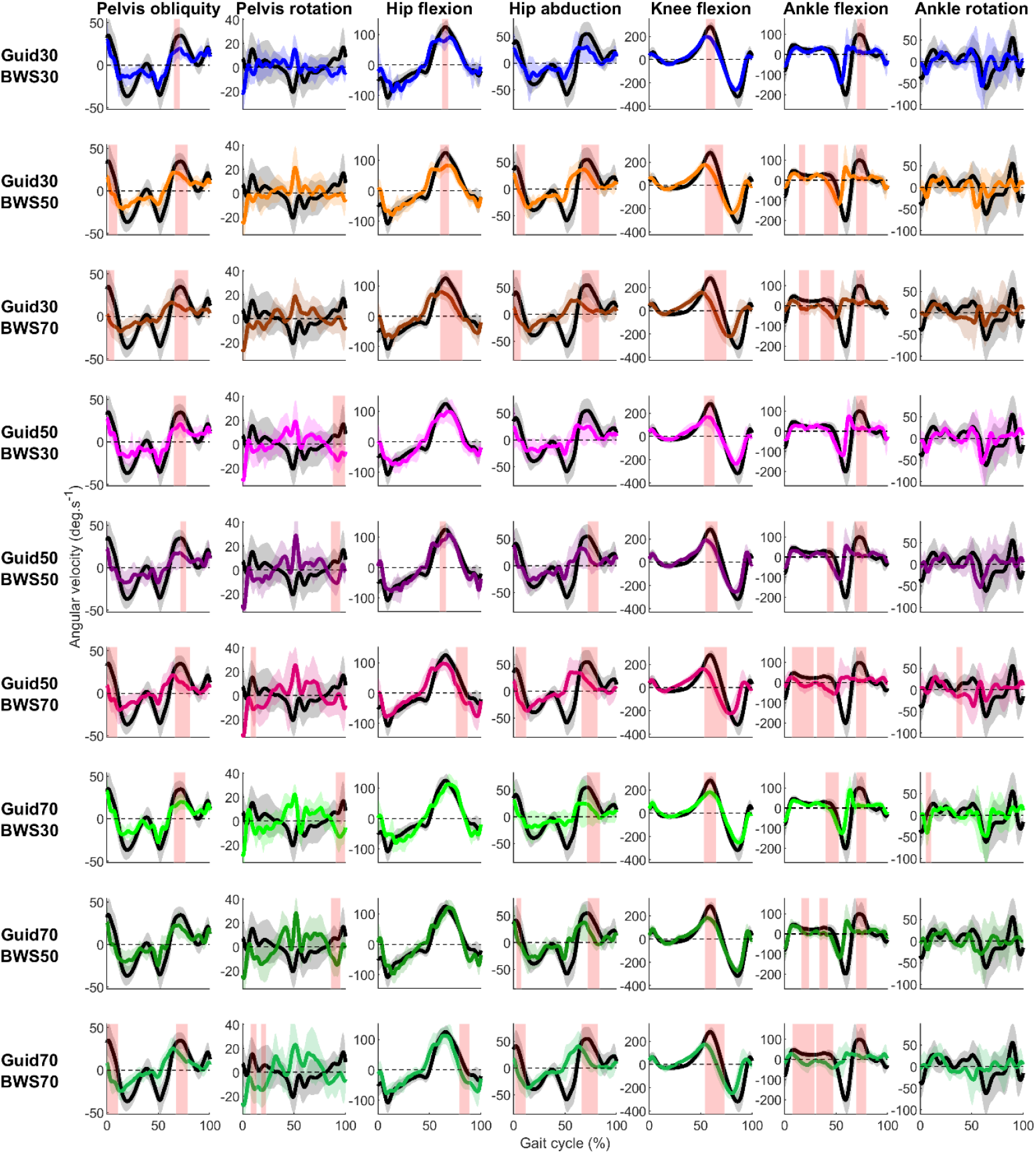
Participant’s mean (solid lines) ± standard deviation (shaded zone) joint angular velocities during the Treadmill (black) and Lokomat (colored) conditions throughout the gait cycle for each degree of freedom (column) and Lokomat setting (row). Light red shaded zones show a significant difference between the Treadmill and Lokomat condition (FDR corrected *p*-values are reported in **Table S1**).

**Figure 5.**
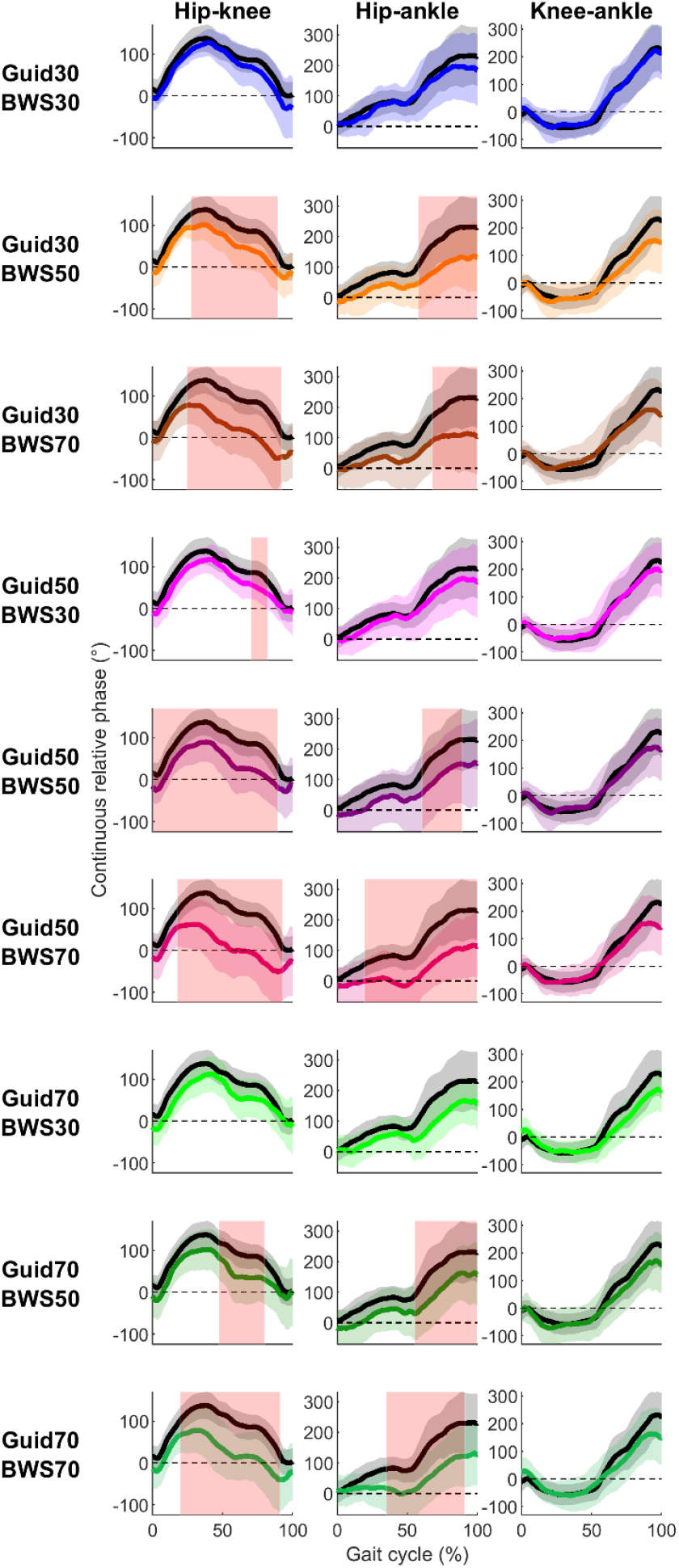
Participant’s mean (solid lines) ± standard deviation (shaded zone) CRP during the Treadmill (black) and Lokomat (colored) conditions throughout the gait cycle for each joint couple (column) and Lokomat setting (row). Light red shaded zones correspond to significant difference between the Treadmill and Lokomat condition (FDR corrected *p*-values are reported in **Table S1**).

### 3.1 Joint angles

In the <u>sagittal plane</u>, the Lokomat condition presented few significant differences from the Treadmill condition (**Figure 2**). For the hip flexion, there was no difference reported, and correlations were strong (0.88≤R≤0.97) (**Figure 6**). Knee and ankle showed significantly smaller flexion in the Lokomat than the Treadmill condition with moderate to strong (0.65≤R≤0.98) and low to moderate correlations (0.10≤R≤0.71), respectively. Significant differences lasted less than 20% of the gait cycle and occurred during the swing phase and the end of the stance phase for the knee and the ankle, respectively.

**Figure 6.**
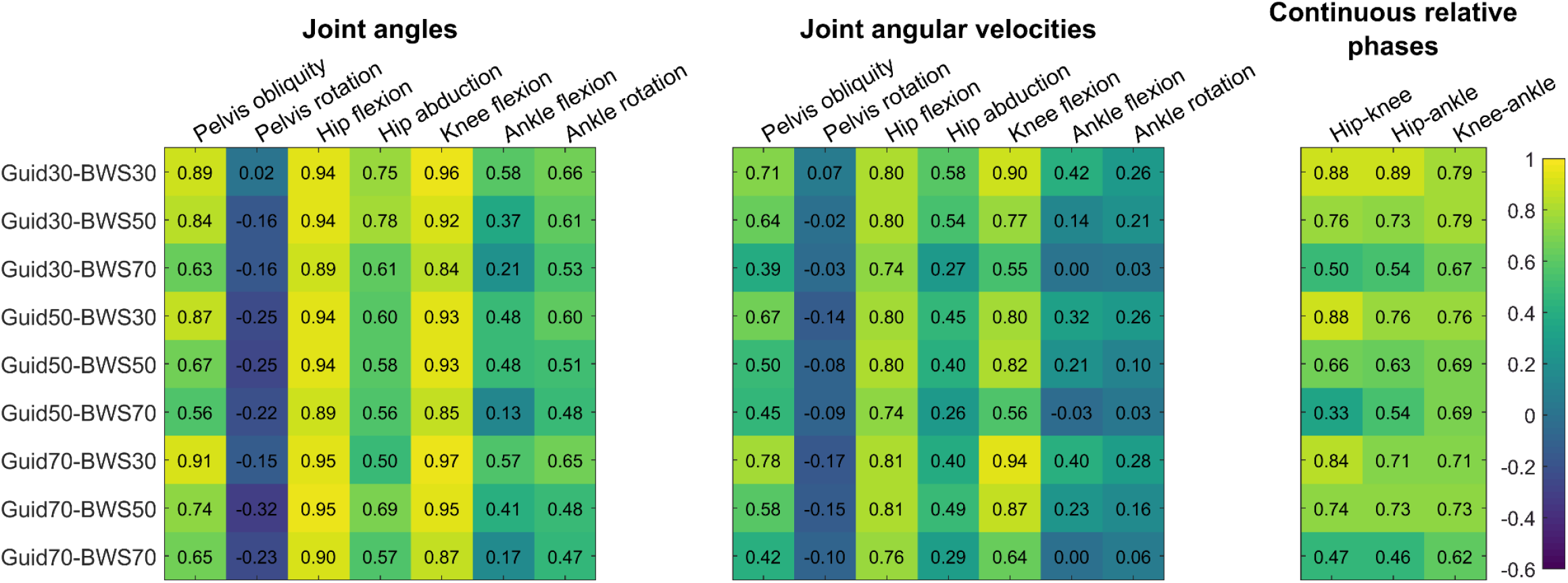
Heat map representation of correlation coefficients between the Treadmill and each Lokomat condition for joint angles and angular velocities and continuous relative phases. Numbers represent correlation coefficients.

In the <u>coronal and transverse planes</u>, the Lokomat condition was characterized by systematic smaller pelvis obliquity and hip adduction and smaller pelvis and ankle internal rotation than the Treadmill condition (**Figure 2**). Significant differences were reported during the stance phase for the pelvis obliquity, pelvis rotation, and hip adduction and around the toe-off period for the ankle internal rotation. Correlations were moderate to strong for the pelvis obliquity (0.56≤R≤0.93), low for the pelvis rotation (−0.32≤R≤0.02), moderate for the hip abduction (0.43≤R≤0.73), and for the ankle rotation low to moderate (0.18≤R≤0.56) (**Figure 6**).

### 3.2 Joint angular velocities

All Lokomat settings significantly reduced participants’ stride rates compared to Treadmill (**Figure 3**). In the <u>sagittal plane</u>, 24 out-of the 27 comparisons made between the Lokomat and the Treadmill conditions showed significantly slower velocities for the hip, knee, and ankle flexion. For the hip and knee, differences were reported around the toe-off period (**Figure 4**) with moderate to strong correlations (0.74≤R≤0.88 and 0.55≤R≤0.94, respectively). For the ankle, differences were reported in the early swing phase for Lokomat conditions with 30% of BWS and during the stance phase for Lokomat conditions with 50% and 70% of BWS. Correlations for ankle flexion were low to moderate (0.00≤R≤0.42).

In the <u>coronal and transverse planes</u>, pelvis obliquity and hip abduction were significantly slower during the swing phase for 15 out-of 18 comparisons made between the Lokomat and the Treadmill conditions (**Figure 5**). Correlations were moderate to strong for pelvis obliquity (0.396≤R≤0.78) and low to moderate for hip abduction (0.26≤R≤0.58) (**Figure 6**). Concerning pelvis rotation, short-lasting significant differences were reported around the heel strike period between the Lokomat and the Treadmill condition for all Lokomat conditions with 50% and 70% of BWS. Correlations were low (−0.17≤R≤0.07).

### 3.3 Continuous relative phases

For hip-knee and hip-ankle CRP, significant differences were reported between Lokomat and Treadmill condition for 13 out of 18 Lokomat conditions. Overall, these differences started at the early/mid-stance phase and lasted until the late swing phase (**Figure 5**). Moderate to strong correlations were observed between Lokomat and Treadmill conditions in terms of CRP between hip-knee (0.67≤R≤0.91), hip-ankle (0.66≤R≤0.85), and knee-ankle (0.90≤R≤0.95) (**Figure 6**).

## 4. Discussion

This study aimed to compare the robotic-assisted Lokomat gait with different levels of BWS and Guidance with a non-assisted Treadmill gait in a healthy population. Joint angles, angular velocities, and coordination were assessed during Lokomat and Treadmill conditions.

### 3.4 Sagittal plane kinematic

Regarding joint angles, hip and knee flexion showed strong similarity between the Treadmill and the Lokomat conditions with significant differences only for knee flexion angle during 2 out of 9 Lokomat settings, namely, BWS set at 70% combined with Guidance and 30% and 50%. This result agrees with the visual inspection previously made by Hidler et al., (2008) that the Lokomat, at least partially, mimics normative gait. Concerning ankle joint, although no significant differences were found between Lokomat and Treadmill conditions, weak correlations, especially when 70% of the participants’ body-weight was supported, were reported suggesting large inter-participants variability at this degree-of-freedom. This result may be explained by the ankle actuators of the Lokomat made by springs that provide less control on joint kinematics than motors actuating hips and knees.

More unexpected results were obtained concerning angular velocity and inter-joint coordination. Indeed, although the Lokomat drives participants’ legs in the sagittal plane, hip and knee flexion/extension peak velocity around the toe-off period and hip-knee and hip-ankle coordinations were altered. These results may agree with increased knee flexor and extensor muscle activation levels during the Lokomat gait compared to the Treadmill gait (Cherni et al., 2021). Indeed, the Lokomat perturbed lower limb dynamic pattern, which is known to increase muscle activations (Barthélemy et al., 2012). In addition, although walking speed was constant between Lokomat and Treadmill conditions, Lokomat gait was characterized by a slower stride rates and then a lower angular velocity at the hip, knee and ankle. This results is in line with the results of Umberger and Martin, (2007) that showed an increase in peak hip and knee angular velocities as the stride rate increased in healthy people walking at a same speed. Although these results indicate that the Lokomat partially replicates normal gait in the sagittal plane, improvements of hip and knee actuator controllers may be required to increase its clinical efficiency.

### 3.5 Coronal and transverse plane kinematics

As expected, some differences between the Lokomat and Treadmill conditions were mainly found in the coronal and transverse planes. A reduced pelvis obliquity and hip adduction as well as an inversed pelvis rotation were observed during the stance phase when participants walked in the Lokomat compared to the Treadmill. This result may be a direct consequence of the reduced pelvis translations. Indeed, the Lokomat limits pelvis lateral displacement, which has been shown to reduce its obliquity during the stance phase (Hidler et al., 2008). Moreover, as the cuffs are tightened to prevent leg lateral movements, the Lokomat limits hip adduction.

As observed in the sagittal plane, differences between the Lokomat and Treadmill gaits increased with the BWS level. This interpretation is supported by the concomitant absence or short-lasting significant decrease of pelvis obliquity and hip adduction during experimental conditions with 30% of BWS. Indeed, although the Lokomat drives the participants’ legs in the sagittal plane only, strong similarity was reported between Lokomat and Treadmill gaits for pelvis obliquity and hip abduction angle as well as pelvis obliquity angular velocity. These motions may come from the compression of the hip pads surrounding the greater trochanters, as already evidenced (Swinnen et al., 2015b). This result is observed for lesser BWS as this condition increases ground reaction forces so that larger pelvis medio-lateral contact force may be applied against pads at the greater trochanters that may increase their compression and favor pelvis obliquity. The optional Lokomat FreeD module, which enables pelvis lateral translation coupled with transverse rotation for a more normative gait pattern, that has already been shown to increase trunk lateral displacement (Aurich-Schuler et al., 2019) may contribute to preserving more normative gait pattern during Lokomat gait with BWS larger than 30%.

### 3.6 Clinical implications

Although in clinical practice the objective of the Lokomat may not be to exactly reproduce normative gait patterns since patients may never be able to walk typically without assistance (Hidler et al., 2008), some significant discrepancies between the Lokomat and the Treadmill gaits may limit the clinical impact of this training modality. In the <u>sagittal plane</u>, the hip, knee, and ankle angular velocities were slower in the Lokomat than the Treadmill. Because of its relationship with the gait dynamics, joint angular velocities are important to be considered (Granata et al., 2000). Indeed, joint angular velocities are in general reduced in patients who have spasticity due to spinal cord injury (Krawetz and Nance, 1996). Thus, the Lokomat may not be the optimal modality if the clinical objective is to increase joint excursion in spastic patients. Regarding joint coordination, walking in the Lokomat altered hip-knee and hip-ankle coordination. However, Lokomat accurately reproduced similar knee-ankle coordination to the Treadmill condition. Although kinetic data on joint torque would be required to ascertain this interpretation, the Lokomat may adequately reproduce the plantarflexion-knee extension couple, which plays an important role in knee control during gait (Brunner and Rutz, 2013). For example, when the plantarflexion–knee extension couple disappears, a crouch gait occurs such as in cerebral palsy patients (Brunner and Rutz, 2013). Regarding the <u>coronal plane</u>, walking in the Lokomat reduced pelvis obliquity and hip adduction. The hip adduction plays a crucial role in controlling the mediolateral displacement of the body center of mass and thus in dynamic balance (MacKinnon and Winter, 1993; Saunders et al., 1953). The latter is often significantly impaired in people with neurologic disorders such as acquired brain injury or multiple sclerosis (Kaufman et al., 2006; Morrison et al., 2016). A combination of 30% of BWS with 30% of Guidance seems to be the most suitable to minimize the discrepancy between the Lokomat and Treadmill in terms of hip adduction and could be relevant for patients with dynamic balance disorders.

Although there is some evidence from clinical trials that the Lokomat settings such as BWS and Guidance are important to maximize patient benefits from this robotic technology (Lefmann et al., 2017), there were currently no guidelines for best practice. This lack of guidelines and the difficulty in defining optimal settings present a major constraint in achieving the expected goals following Lokomat training. Based on the present results, and in agreement with previous findings on trunk and pelvis kinematics during Lokomat gait (Swinnen et al., 2015a, 2015b), the BWS, but less the Guidance, tended to mitigate the effect of Lokomat on lower limb kinematics. Indeed, gait during the conditions with 30% of BWS presented strong similarity with Treadmill gait regardless of the Guidance level. These results are also in agreement with previous studies showing that high levels of BWS, as well as low Guidance, result in typically low muscle activation as well as an alteration of lower limb muscle coordination (Cherni et al., 2021; Kammen et al., 2014).

### 3.7 Limitations

There are some limitations in this study that need to be considered. Firstly, only healthy young adults were evaluated, while the Lokomat has been designed for people with gait disorders. Nevertheless, it was impossible to perform similar comparison with patients since they would not be able to perform a non-assisted gait on Treadmill. Secondly, the Treadmill condition was performed after the Lokomat conditions, which does not allow us to completely exclude an aftereffect of Lokomat.

## 5. Conclusion

Based on the present findings, the Lokomat allows to partially reproduce the normative gait pattern. Although the Lokomat reproduces well enough the kinematics in the sagittal plane, walking in the Lokomat results in an altered pelvis rotation, a reduced pelvis obliquity, and hip adduction during the stance phase compared to the Treadmill. The results of the present study suggest that therapists should be aware that alterations in the gait pattern may occur for certain combinations of Lokomat settings. Overall, we recommend a BWS at 30% to get closer to the normative gait patterns.

## Data Availability

All data produced in the present study are available upon reasonable request to the authors

## Acknowledgements

The authors acknowledge the FRQS, GRSTB and the Méditis (NSERC) program that funded this research. The authors also acknowledge Veronica Cuevas Villanueva, Émilie De La Sablonnière-Griffin, and Yalpe Nismou for their assistance in data collection.

## Conflict of interest

The authors state that there are no conflicts of interest.

## Notes

### Competing Interest Statement

The authors have declared no competing interest.

### Funding Statement

None

### Author Declarations

Ethics Committee of Sainte-Justine Hospital (#4049).

