## Supplemental Table 1 for "Effects of Robotic-Assisted Gait at Different Levels of Guidance and Body Weight Support on Lower Limb Joint Angles, Angular Velocity, and Inter-Joint Coordination": Supplementary material.docx

**Table S1.** Intervals in the percentage of the gait cycle and FDR corrected *p*-values in parenthesis of significant differences shown by the statistical parametric mapping procedure between the Treadmill and each Lokomat condition

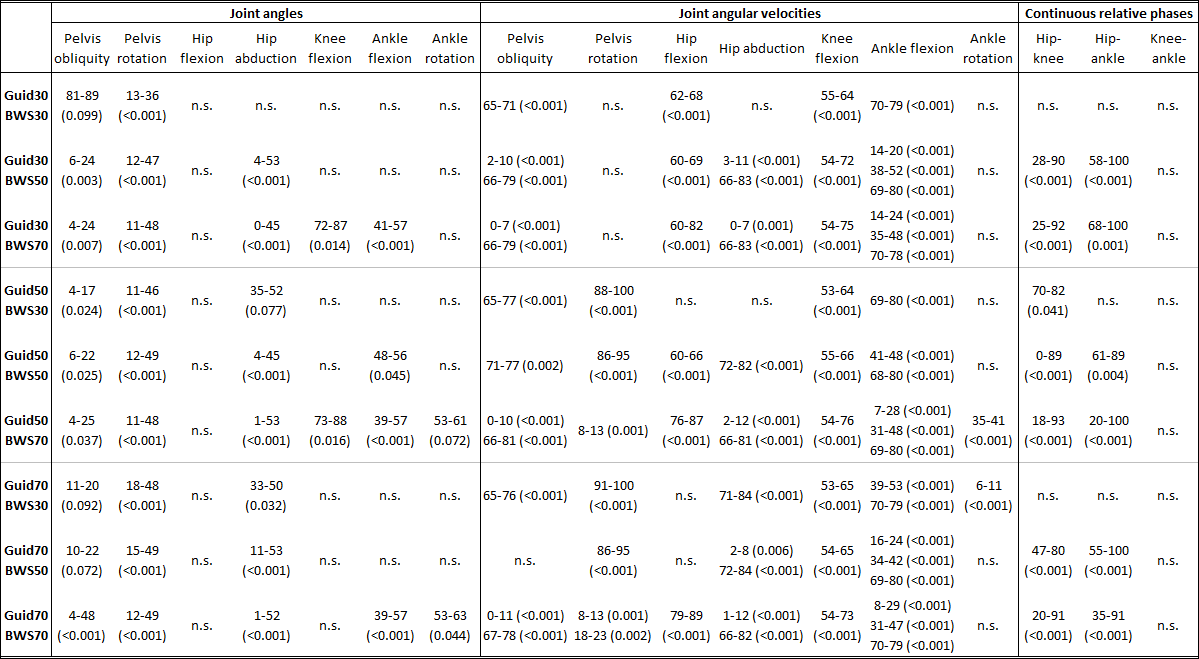
